# Risk Factors for Post-Traumatic Stress Disorder (PTSD) in COVID Survivors: A Cross-Sectional Study

**DOI:** 10.1101/2023.08.23.23294470

**Authors:** Surakshaya Dhakal, Ram Prasad Khatiwada

## Abstract

The study aimed to investigate the impact of demographic, socio-economic, health, and lifestyle variables on the development of PTSD symptoms in COVID survivors. The study used a cross-sectional design, and data were collected via a standard set of questionnaires from 228 COVID survivors, who required oxygen support and were admitted to Damak COVID hospital from April to October 2021.

Descriptive statistics such as frequency and percentage were used to summarize the data and inferential statistics such as chi-square test, Fisher’s exact test, and Binary logistic regression were used to analyze the data and to infer the overall result from the taken sample. The study found that only three variables, i.e., gender, diabetes, and chronic obstructive pulmonary disorder (COPD), were significant factors that posed a higher threat of PTSD in COVID survivors. Additionally, the study uses model adequacy tests such as Pseudo R2 test, Reliability test and Hosmer and Lemeshow test to validate the model fitted.

The study found that only three variables had significant impact PTSD symptoms in COVID survivors. Male patients were more likely to have PTSD symptoms than female patients. The presence of diabetes before or after the infection increased the risk of PTSD. The patients with high blood pressure before COVID and those who developed chronic obstructive pulmonary disorder (COPD) after COVID were more likely to experience PTSD symptoms. The study provides valuable information on the risk factors for developing PTSD symptoms in COVID survivors. This study can contribute to the understanding and growing body of research on the psychological impact of COVID and help healthcare professionals identify patients who are at risk of developing PTSD and provide them with appropriate interventions to prevent or treat PTSD.

## INTRODUCTION

### Background of Study

In December 2019, pneumonia cases of unknown cause were reported in Wuhan, China. Later, it was identified as a virus called SARS-CoV-2, causing severe acute respiratory syndrome [1].On January 30, 2020, the WHO declared it a global Public Health Emergency, recognizing it as a major epidemic [2]. Coronaviruses are RNA viruses found in birds and mammals, including humans. They can cause respiratory, enteric, and hepatic diseases. While four coronaviruses only result in common cold-like symptoms in immunocompromised individuals, two strains—SARS-CoV and MERS-CoV—have caused severe and fatal illnesses [3].

New data shows that more than 1 million people in Nepal have contracted SARSCoV-2, and the recovery rate is over 98.5% [4]. Some survivors bounced back quickly, but others experienced lasting physical and mental effects. The World Health Organization (WHO) estimates that it can take six weeks or longer to recover from a COVID infection [5]. Various factors, including age, gender, and underlying health conditions, can affect the rate and duration of patient recovery. Older males with chronic pulmonary conditions who needed intranasal oxygen supplementation had a longer recovery time [6]. A study published by the Centers for Disease Control and Prevention (CDC) in the Morbidity and Mortality Weekly Report revealed that even patients with milder illness can experience prolonged symptoms. After 2-3 weeks of recovery, more than 35% of patients reported persistent issues like cough, fatigue, or shortness of breath [7].

Throughout history, human civilization has experienced numerous epidemics, including the ongoing one, and few phenomena have had as profound an impact on societies and culture as disease outbreaks. Major pandemics such as the Spanish flu (1918-1920), the HIV pandemic of the late nineteenth century, and the Swine flu pandemic of the early twentieth century have greatly affected human society. They have devastated communities, influenced the outcomes of wars, and even led to the extinction of entire populations. However, these epidemics have also stimulated scientific progress and advancements, particularly in the fields of public health and medicine [8]. In times of trauma, such as natural disasters, sexual harassment, accidents, war, medical emergencies, or the loss of loved ones, humans experience a range of psychological manifestations. While some of these manifestations are expected and normal, others can manifest as varying degrees of mental health issues. Immediate problems often include depression and acute stress reactions, while delayed forms of psychological manifestations commonly include pathological grief, depression, adjustment disorders, post-traumatic stress disorder, substance abuse, and psychosomatic disorders [9].

Posttraumatic Stress Disorder (PTSD) is a common psychological condition resulting from trauma, characterized by recurring mental and physical distress triggered by memories of a past traumatic event. Individuals with PTSD intentionally avoid thoughts, memories, and discussions about the event [10]. PTSD is marked by intense fear, persistent re-experiencing, avoidance, and hyperarousal symptoms that can last for months or even years after the trauma. Multiple factors contribute to the likelihood of developing PTSD, with genetic factors accounting for about 30-40% of the risk [11]. Previous experiences, including adult and childhood trauma, also influence the susceptibility to PTSD. Traumatic events encompass natural disasters, disease outbreaks, substance abuse, war, domestic violence, and more [12] [13] [14]. Survivors of COVID-19 may experience psychological effects due to the life-threatening nature of the infection. Physical trauma survivors may suffer from nightmares, flashbacks, intrusive thoughts, and behavioral changes such as hypervigilance, hyperarousal, avoidance, sleep difficulties, and angry outbursts [15]. The widespread availability of social media news contributes to anxiety but does not directly impact mental health. However, it has been shown to contribute to Secondary Traumatic Stress (STS), despair, and anxiety [16]. Health Care Workers (HCWs) and COVID-19 patients have also reported symptoms of PTSD and post-traumatic stress [17] [18].

The global attention on COVID-19 has overlooked its unknown physiological effects. While the physical harm caused by the pandemic is widely discussed, its psychological impact remains overlooked. Each successive mutant strain of the virus is believed to be more contagious than its predecessor. Even after two years since its emergence, COVID-19 continues to affect the entire planet, prompting efforts to address economic inequality and minimize fatalities. However, the psychological trauma inflicted by the pandemic has received little attention, particularly in developing and disadvantaged countries.

Nepal, with a limited public health budget [19], struggled to handle the surge of patients during the pandemic, leading to a critical shortage of oxygen [20]. Despite a relatively low rate of serious infections, hospitals were overcrowded, resulting in many patients being unable to receive basic care, such as artificial oxygen. The focus of the authorities was primarily on controlling the pandemic, with little attention given to psychological well-being. Mental health facilities in Nepal are primarily located in major cities, with a scarcity of professionals, including 0.22 psychiatrists and 0.06 psychologists per 100,000 population. Moreover, the country has only 18 outpatient mental health facilities, indicating a lack of efforts to address and understand the psychological needs of patients [21]. Although a few public and private organizations have contributed to psychology, there is a significant dearth of services and research projects outside the capital city [22].

This study examines PTSD prevalence among severe COVID patients who required oxygen support during their hospital stay at Damak COVID hospital. The hospital, built by Damak Municipality as a primary care facility for COVID-19 patients, now serves as a vital treatment center for Damak and neighboring areas. Located near the Indo-Nepal border, Damak is a large and populous municipality that attracts travelers from the eastern and southern frontiers, leading to a higher incidence of reported cases and increased patient visits to the hospital.

## MATERIALS AND METHODS

### Data and study area

The study focused on discharged patients from Damak COVID Hospital, which situated in the northern part of Damak Municipality near Ratuwa River. The patients were from the local community covering 75.42 sq. km and surrounding areas such as Urlabari (west), Lakhanpur (east), Gauradaha (south), and Chulachuli (north). The researcher personally visited the participants at their homes and asked them to complete the questionnaire.

The study used a cross-sectional study design. It included inferential and descriptive findings. Primary information was collected through in-person interviews. To assist participants who struggled with English, the questionnaire was first developed in English and then translated into Nepali.

According to Damak COVID Hospital, a total of 589 COVID-19 positive patients were admitted from April 14, 2021 to October 16, 2021. The data was collected in collaboration with the Damak Municipality office. Out of these patients, 338 were male and 251 were female. The study did not include patients who were transferred or deceased. Also, 28 patients were referred, and among them, 4 passed away.

Data were collected through self-administered interviews using a PTSD questionnaire set, including the PTSD checklist-Civilian (PCL-C) [23]. When tested in Nepal, PTSD checklist-Civilian (PCL-C) demonstrated positive psychometric qualities in terms of internal consistency, test-retest reliability, convergent validity, and discriminant validity. Participants’ health and lifestyle variables were recorded based on their own responses. The study included 557 patients who tested positive for COVID-19 and required artificial oxygen after being hospitalized for at least one day. The sample was selected using simple random sampling, and data collection took place from December 2022 to January 2023.

A sample size of 228 was determined using the Yamane formula, considering a 95% confidence level, a 5% margin of error, and a 50% population proportion [24]. The study collected data from the records of the Damak COVID hospital to determine the sample size based on a finite population size.

### Variables

The dependent variable of this study is dichotomous i.e. either prevalence of PTSD or non-prevalence of PTSD. Different category variables are chosen as explanatory variables by evaluating the pertinent literature. Further, explanatory variables were classified into socio-demographic, health-related, and life-style related variables. Socio-demographic variables include age category, gender, ethnicity, employment status, marital status. Similarly, health related variables Presence of Diabetes, High Bp, COPD, Cardiovascular disorder, Kidney disorder, and Psychological disorder are studied. Life-style related variables; smoking status, alcohol-consuming status, habits of doing yoga and whether the respondent is vegetarian or not considered in the study. Both health-related and life-style related variables are studied directly by the answer given by the respondent without any medical supervision or observation. They are categorized as they have them from before the COVID infection, only after the infection or do not have till now.

The purpose of this study is to understand whether people have PTSD (Post-Traumatic Stress Disorder) or not. We looked at different factors that might explain this. We chose different categories to explain why this is the case, based on what we found in relevant research. These categories include socio-demographic factors (like age, gender, ethnicity, employment status, and marital status), health-related factors (such as diabetes, high blood pressure, COPD, cardiovascular disorder, kidney disorder, and psychological disorder), and lifestyle factors (like smoking, alcohol consumption, yoga habits, and whether the person is vegetarian or not). We collected this information directly from the respondents without any medical supervision or observation. We categorized the variables based on whether they had these factors before, only after the COVID infection, or if they don’t have them at all.

The study used the PCL-C (Posttraumatic Stress Disorder Checklist) to identify PTSD. PCL -C is a standardized self-report scale with 17 items that correspond to key PTSD symptoms. Two versions of the PCL exist: PCL-M for military-related PTSD and PCL-C for any traumatic event. In this study, the PCL-C version was used to identify PTSD in post-COVID patients. Scoring of the PCL-C version was done by adding all the items for a total severity score or response categories 3–5 (Moderately or above) were treated as symptomatic and responses 1–2 (below moderately) were treated as non-symptomatic. Then the DSM criteria were used for the diagnosis where, Symptomatic response to at least 1 “B” item (Questions 1–5), symptomatic response to at least 3 “C” items (Questions 6–12), and symptomatic response to at least 2 “D” items (Questions 13–17) are considered as symptomatic [25].

### Measure of reliability

Cronbach’s alpha is a valuable measure for internal consistency. In the case of Likert scale questionnaires about PTSD, the reliability measure obtained from pilot surveys was 0.88. A value above 0.69 is considered acceptable for Cronbach’s alpha [26] as given in Table 1, and 0.88 falls within the good range. This indicates that the scales used are reliable.

**Table 1:** Range of Cronbach’s alpha coefficients and their reliability levels.

| S.N | Coefficient of Cronbach’s Alpha | Reliability test |
| --- | --- | --- |
| 1 | More than 0.90 | Excellent |
| 2 | 0.80-0.89 | Good |
| 3 | 0.70-0.79 | Acceptable |
| 4 | 0.6-.69 | Questionable |
| 5 | 0.5-0.59 | Poor |
| 6 | Less than 0.59 | Unacceptable |

### Data Analysis

The study’s findings were analyzed using descriptive and inferential methods. Descriptive analysis involved calculating frequencies and percentages of participant attributes. In the inferential section, several model adequacy tests such as Pseudo-R square, Hosmer-Lemeshow test, and accuracy check for the dependent variable were conducted to determine the overall significance of the model coefficient. Additionally, the chi-square test, Fisher Freeman exact test, binary logistic regression analysis, and odds ratio were calculated.

Binary logistic regression is a type of linear regression model used when the outcome variable is dichotomous, taking values of either 0 or 1, and is influenced by one or more continuous or categorical explanatory variables. In this study, the explained variable is PTSD, and it is determined based on the values of a set of predictor variables. The dependent variable PTSD is categorized as: Y_i_= 0, indicating not having PTSD, and Y_i_ = 1, indicating having PTSD. The independent variables, denoted as X_1_, X_2_,……., X_k_, can be discrete or continuous or a combination of both. The mathematical model for binary logistic regression is expressed as follows:

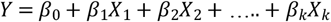

Its specific form is 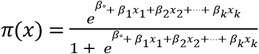

The logit transformation in terms of g(x) is

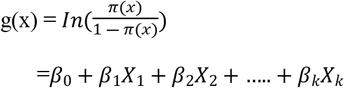

where π(x) is the probability that the event occurs π (y=1)

g(x) is the logit transformation and also called logit model.

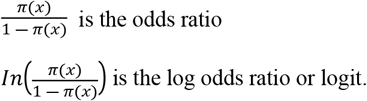

### Model Adequacy Test

Pseudo R square is used to assess the strength of logistic regression. While the measure of correlation is not sufficient for evaluating the accuracy of error associated with the model in binary logistic regression, this study utilized Cox and Snell R-square and Nagelkreke R-square to quantify the extent to which the independent variables explain the variation in dependent variables. Additionally, the Homser-Lemeshow test was conducted to measure the goodness of fit statistics of model fitting. A smaller difference between observed and predicted classification, or a large p-value (greater than 0.05), indicates a better fit for the model.

## RESULTS

Out of 228 respondents, 92 (40.4%) showed symptoms of PTSD, while 136 (59.6%) did not report such symptoms. The participants were categorized based on demographic variables. Of the total 228 participants, 124 (54.4%) were male, and 104 (45.6%) were female. The study classified the participants into five groups: Janajati (43.9%, n=100), Brahmin (29.8%, n=68), Chhetri (15.4%, n=35), Dalit (6.6%, n=15), and others (3.1%, n=7). Regarding marital status, the participants were divided as follows: married 188 (82.5%), unmarried 24 (10.5%), separated 2 (0.9%), divorced 2 (0.9%), and widowed 12 (5.2%). Based on age, six categories were created. The age group of 40-50 years had the highest number of participants at 28.1% (n=64), followed by the age group of 30-40 years at 27.6% (n=64). The age group of 50-60 years had 39 (17.1%) participants, the age group of 20-30 years had 34 (14.9%) participants, and the age group above 60 years had 28 (12.3%) participants. No participants were below the age of 20 years. This may be due to younger individuals having a lower likelihood of severe COVID infections requiring hospitalization or oxygen support.

Another important variable associated with the occurrence of PTSD symptoms was socioeconomic status. Socioeconomic status was found to be an important factor associated with PTSD symptoms. The study examined education level and employment status. Results showed that 11.4% of participants were illiterate, 21.9% were literate but not enrolled in any educational institute, 2.6% had primary level education, 6.1% had lower secondary education, 26.8% had secondary level education, 21.1% had higher secondary education, and 10.1% had a bachelor’s degree or higher.

The study looked at how having a job could affect the number of people with PTSD. They included whether someone was employed or not as a factor. Out of all the people in the study, 55.7% (n=127) had a job, while 44.3% (n=101) did not have a job.

The study did not directly identify health-related variables through clinical means. Instead, questionnaires were utilized to gather information on the participants’ health history. The study examined three categories of health-related variables: 1) health conditions that emerged solely after the infection, 2) health conditions present both before and after the infection and 3) the absence of any such health condition before and after the infection.

Regarding diabetes, 15.8% (n=36) had the condition both before and after COVID, while 6.6% (n=15) developed it after the infection. The majority, 77.6% (n=177), did not have diabetes before or after the infection. For kidney disorders, only 1.3% (n=3) had it before COVID, while 3.5% (n=8) acquired it after the infection. The majority, 95.2% (n=217), did not have kidney problems at the time of the study. In terms of cardiovascular disease, 0.4% (n=1) had it before infection, while 5.7% (n=13) had it after the infection. The majority, 93.9% (n=214), did not have the condition. Regarding obesity, 99.1% (n=226) of participants did not have the problem, 0.4% (n=1) developed it after COVID, and 0.4% (n=1) had it before.

Another important health variable considered was COPD. It was present in 1.8% (n=4) of participants before infection, while 17.1% (n=39) seemed to have acquired it following the infection. The majority, 81.1% (n=185), did not have COPD. Another variable, Psychological disorder is frequently overlooked in our country, and there is limited awareness about them. As a result, 99.6% (n=227) of participants responded that they did not have any psychological disorder till the date of the study. Only 1 (0.4%) participant responded that a psychological disorder was acquired after COVID infection.

The study classified lifestyle-related factors into four groups: (1) individuals who adopted specific habits only after contracting COVID-19, (2) participants who discontinued their habits after getting infected, (3) individuals who maintained their habits both before and after the infection until the day of the study, and (4) those who did not engage in the habit before or after the infection.

Regarding smoking, 8.3% (n=19) of respondents smoked before COVID-19 and still do. 6.1% (n=4) quit smoking after getting infected, while the majority of participants (84.6%, n=193) never had the habit. Only 2 (0.9%) started smoking after getting infected. Similar results were found for drinking, with only 2 (0.9%) starting to drink after getting infected, while 9.2% (n=21) stopped the habit. 34 (14.9%) of them used to drink before and still do, while most participants (82.6%, n=176) were never involved in alcohol consumption. Another lifestyle-related variable considered in the study was yoga, with most participants (79.8%, n=170) not interested or involved in practicing yoga. 6.6% (n=14) started practicing yoga after recovering from the infection, while 1.9% (n=4) stopped doing yoga after getting infected. The remaining participants (11.3%, n=24) had been practicing yoga before the infection, and they continued to do so after recovering, being aware of its benefits. Regarding smoking, 8.3% (n=19) of respondents smoked before COVID-19 and still do, while 6.1% (n=4) quit smoking after getting infected. The majority of participants (84.6%, n=193) never had the habit, and only 2 (0.9%) started smoking after getting infected.

The study discovered that certain factors like gender, marital status, employment status, diabetes, cardiovascular disorder, kidney disorder, COPD, high blood pressure, and practicing yoga had noticeable differences in average scores between individuals with and without symptoms of PTSD. However, there were limited data available for Cardiovascular Disorder and Kidney Disorder in some cases, which could have influenced the overall analysis of the odds ratio. Consequently, these variables were excluded from the logistic regression, and certain categories within certain independent variables were combined due to a small number of participants.

Further analysis of the significant variables obtained from a chi-square test revealed the following associations:

- Males had 2.71 times higher odds of the outcome compared to females (95% CI: 1.307-5.636).
- Individuals with diabetes before and after COVID-19 had 1.24 times higher odds of the outcome variable compared to those without diabetes (95% CI: 1.42-8.47).
- Individuals with COPD after COVID-19 had 1.45 times higher odds of the outcome variable compared to those without COPD (95% CI: 1.73-10.42).
- Individuals with high blood pressure before and after COVID-19 had 2.69 times higher odds of the outcome variable compared to those without high blood pressure (95% CI: 1.13-6.41)

The binary logistic regression model identified gender, presence of diabetes, COPD, and high blood pressure as significant factors for the outcome variable. Notably, only one category for diabetes and COPD was found to be significant in the fitted regression model. Table 2 displays the values obtained after model fitting.

**Table 2:**
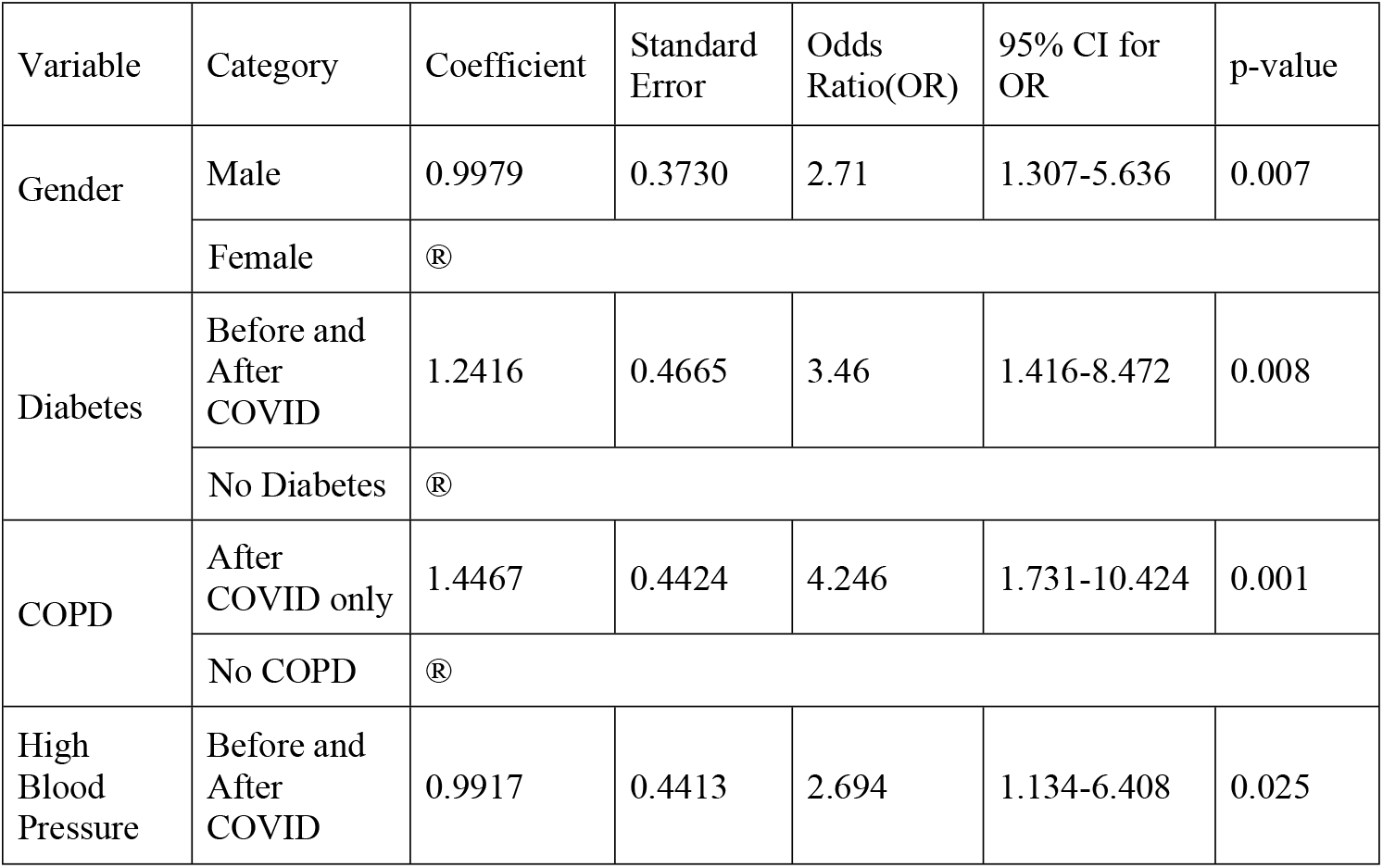
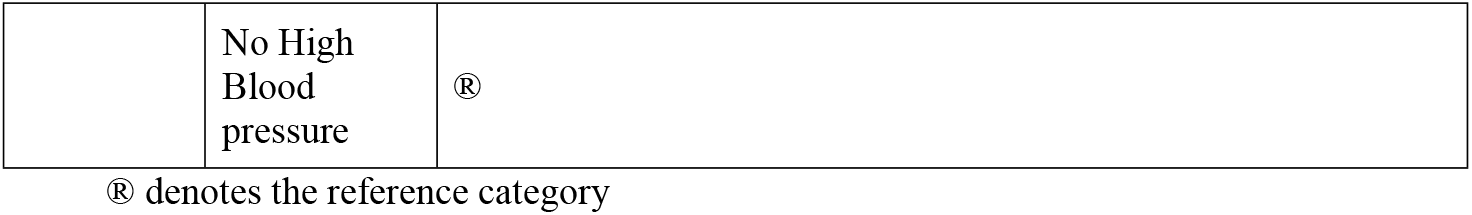
Logistic Regression Model Summary.

| Variable | Category | Coefficient | Standard Error | Odds Ratio(OR) | 95% CI for OR | p-value |
| --- | --- | --- | --- | --- | --- | --- |
| Gender | Male | 0.9979 | 0.3730 | 2.71 | 1.307-5.636 | 0.007 |
|  | Female | ® |  |  |  |  |
| Diabetes | Before and After COVID | 1.2416 | 0.4665 | 3.46 | 1.416-8.472 | 0.008 |
|  | No Diabetes | ® |  |  |  |  |
| COPD | After COVID only | 1.4467 | 0.4424 | 4.246 | 1.731-10.424 | 0.001 |
|  | No COPD | ® |  |  |  |  |
| High Blood Pressure | Before and After COVID | 0.9917 | 0.4413 | 2.694 | 1.134-6.408 | 0.025 |

|  |  |  |
| --- | --- | --- |
|  | No High<br>Blood<br>pressure | ® |

® denotes the reference category

The logistic regression model’s overall fit was evaluated using Cox & Snell R2 statistics and the Hosmer-Lemeshow test [Cox & Snell, Hosmer-Lemeshow]. The results indicated that the model accounted for 24.2% to 32.7% of the variation in the dependent variable (PTSD), depending on whether the Cox and Snell R square or Nagelkerke R square was used as a reference. The Hosmer-Lemeshow test yielded a chi-square value of 6.2604 with 8 degrees of freedom, and the p-value was greater than 0.05, indicating that the model’s goodness of fit was not violated. Additionally, data were assessed for multicollinearity.

### Discussion

COVID leaves deep emotional scars on people with each wave. The study conducted at Damak COVID hospital aimed to identify PTSD in COVID survivors and found significant PTSD symptoms in the target population.

Out of all participants, 40.4% showed a prevalence of PTSD, indicating a high chance of PTSD in the targeted group of people. A similar study conducted by Bo et al. (2021), the prevalence of PTSD in COVID patients was reported to be as high as 96.2% [27]. Another study by Xueyuan Li and colleagues found that 31.6% of Chinese adults experienced PTSD following a COVID infection [28]. These variations in results suggest that the occurrence of PTSD in COVID can be influenced by factors such as geography, society, environment, and lifestyle.

In a study conducted in Nepal by MoHP and NHRC, it was found that 14% of the respondents had anxiety, 7% had depression, and 5% had stress [29]. The frequency of psychological disorders in the Nepal study was much lower compared to the present study. The difference in results could be attributed to the different target populations: the Nepal study included all COVID patients who attended the fever clinic, while the present study focused on COVID survivors who required Oxygen support during hospital treatment. It is evident that patients with more severe conditions experience higher mental pressure and are therefore at a greater risk of developing psychological disorders.

Gender-wise, male COVID survivors exhibited a higher likelihood of displaying PTSD symptoms compared to females. Lei et al. (2021) also found that males had higher odds of developing PTSD than females (OR = 1.484, 95% CI: 1.147 to 1.920) [30]. In contrast to our findings, Wang et al. (2020) reported that the psychological impact of the pandemic was more significant for females than males [31]. Similarly, Martinez et al. (2022) reported a similar situation, with female survivors showing a higher prevalence of PTSD compared to males (OR =1.13, 95% CI: 0.50– 1.76) [32]. The disparity in the development of PTSD between genders across different studies may be attributed to the social and cultural behaviors of the targeted population.

The study showed that people with pre-existing diabetes and those who developed diabetes after COVID are more likely to exhibit symptoms of PTSD compared to those without diabetes. Similarly, individuals with pre-existing high blood pressure (BP) or COPD after the infection are more likely to develop PTSD compared to those without these conditions. However, the presence of high BP after infection and COPD before infection did not show significant associations with PTSD. Thus, the presence or acquisition of conditions such as diabetes and COPD significantly impacts the likelihood of developing PTSD. This finding aligns with previous studies that indicate COVID-19 patients with comorbidities like diabetes, psychological disorders, high BP, obesity, and COPD are more prone to developing PTSD compared to those without these conditions. Previous research by González-Sanguino et al (2020) have identified chronic pulmonary disease as a significant risk factor (OR = 6.03, 95% CI: 1.0–37.1, p = 0.053) for PTSD [33]. Moreover, a web-based survey conducted via the WhatsApp platform reported a higher prevalence of psychological disorders (specifically depression) among COVID-19-infected patients with diabetes mellitus (37.8%), suggesting that diabetic patients are more susceptible to developing PTSD compared to non-diabetic individuals [34]. This result aligns with the fact that diabetic patients typically have a 24% increased risk of developing depressive symptoms even without a COVID-19 infection [35].

## CONCLUSION

In Nepal, there is a scarcity of research focusing on the psychological consequences experienced by individuals who have survived COVID-19. The present study aims to contribute to this knowledge gap by examining the impact of COVID-19 on the mental well-being of survivors. The study specifically investigated the prevalence of post-traumatic stress disorder (PTSD) among individuals admitted to Damak COVID hospital and analyzed the statistical relationship between PTSD symptoms and various factors.

The findings of the study revealed that the prevalence of PTSD among COVID-19 survivors admitted to Damak COVID hospital is moderately high, with approximately 40.4% of the participants exhibiting symptoms of PTSD. Furthermore, the study identified several significant factors associated with the development of PTSD symptoms. Being male, having a diabetic disorder, chronic obstructive pulmonary disease (COPD), and high blood pressure are identified as significant factors contributing to the occurrence of PTSD among COVID-19 survivors.

Overall, this study sheds light on the psychological impact of COVID-19 on survivors in Nepal, filling a crucial gap in the existing research. The identification of significant factors associated with PTSD symptoms provides valuable insights for healthcare professionals and policymakers, enabling them to develop targeted interventions and support systems to address the mental health needs of COVID-19 survivors in the country.

## LIMITATIONS

There were certain limitations in the study due to its scope. Firstly, the medical information provided by the patients could not be verified, as no healthcare professionals were involved. Therefore, all the information regarding past and present disorders relied solely on the patients’ responses.

Additionally, the study was limited to patients admitted during a specific time period due to resource constraints. The data on patients were collected between late 2022 and early 2023, within a year of their hospital admission. It is important to note that over time, the effects of the trauma experienced by the patients may have diminished.

## Data Availability

All relevant data are within the manuscript and its Supporting Information files. However, the personal information including the Name and Contact number of the respondent cannot be made available due to ethical issue as the information were promised to be kept confidential during the data collection procedure.

## Ethical Approval and Respondent’s Consent

In order to conduct the research, ethical approval was obtained from the Institutional Review Committee (IRC) of the Institute of Science and Technology (IOST), Tribhuvan University (TU). Additionally, In accordance with the study protocol, prior informed consent was obtained from all the participating patients to gather relevant information pertaining to them. This ensured that the research adhered to ethical guidelines and respected the rights and privacy of the individuals involved.

## Acknowledgements

The authors wish to sincerely thank the Head of the Department and all the professors from the Central Department of Statistics at Tribhuvan University for their valuable feedback and encouragement during the paper’s completion. Furthermore, the authors would like to acknowledge the support of Damak municipality and Damak COVID Hospital for the research.

## Supporting Information

**S1 Table.**
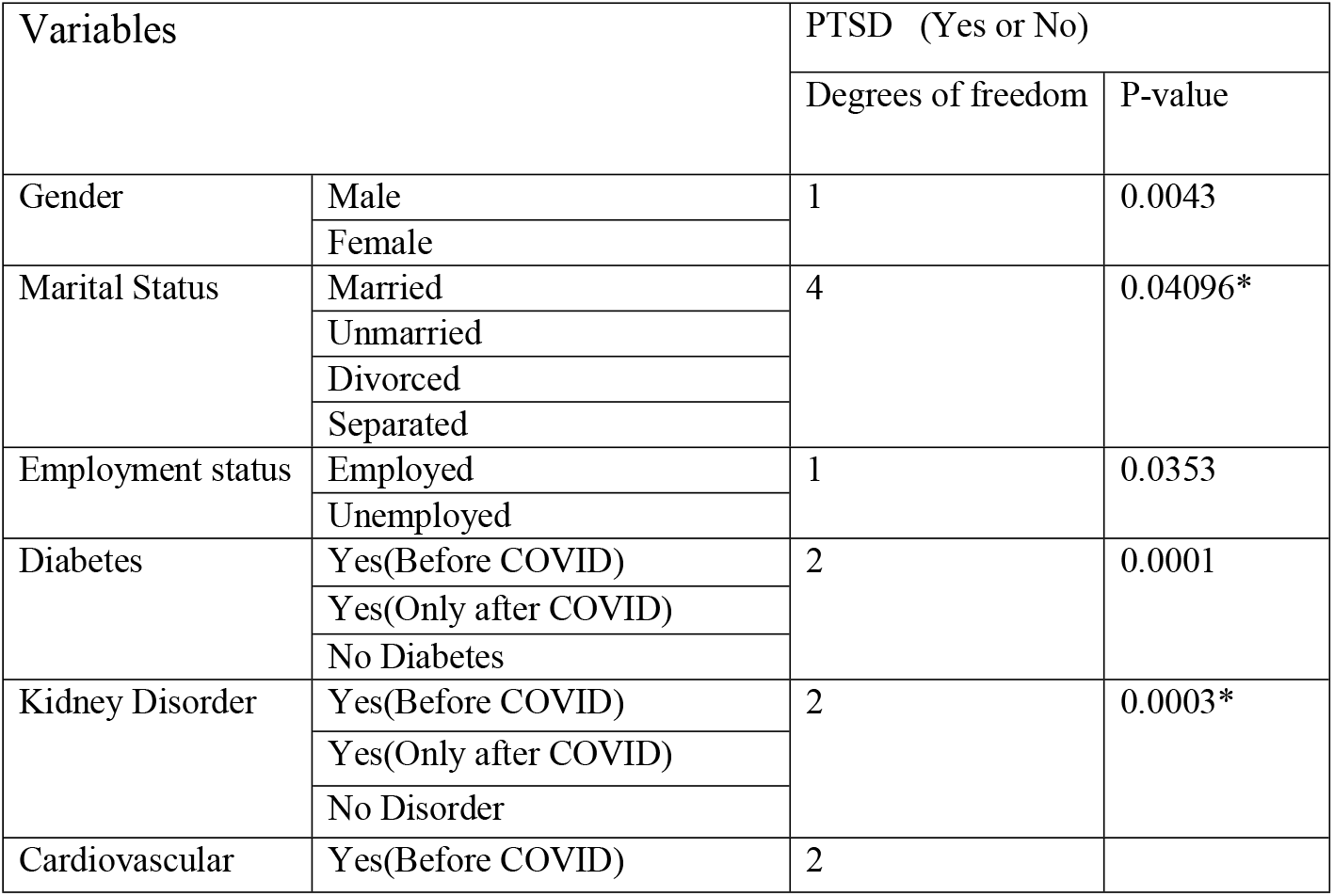
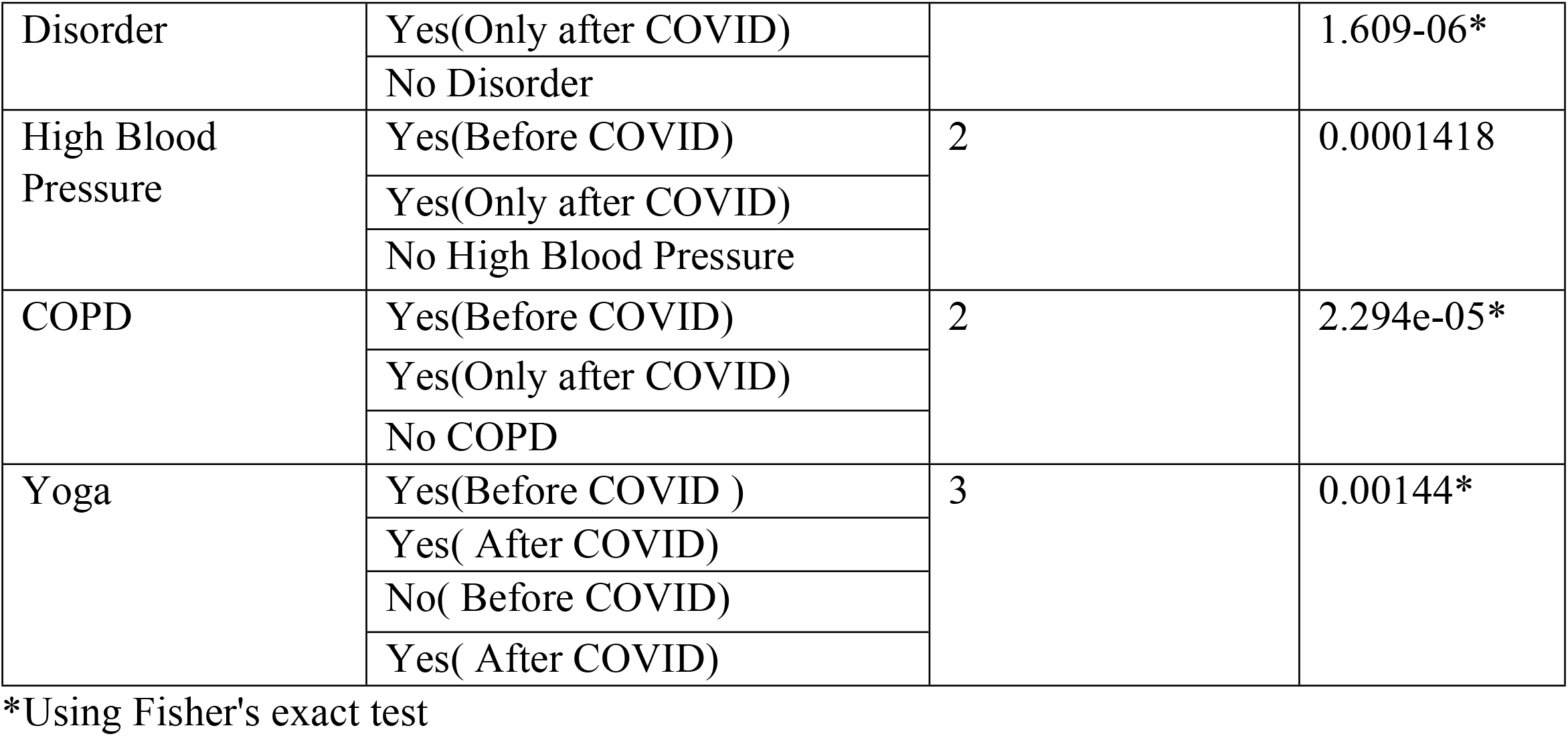
Bivariate analysis of different significant variables in relation to PTSD.

**Fig 1:**
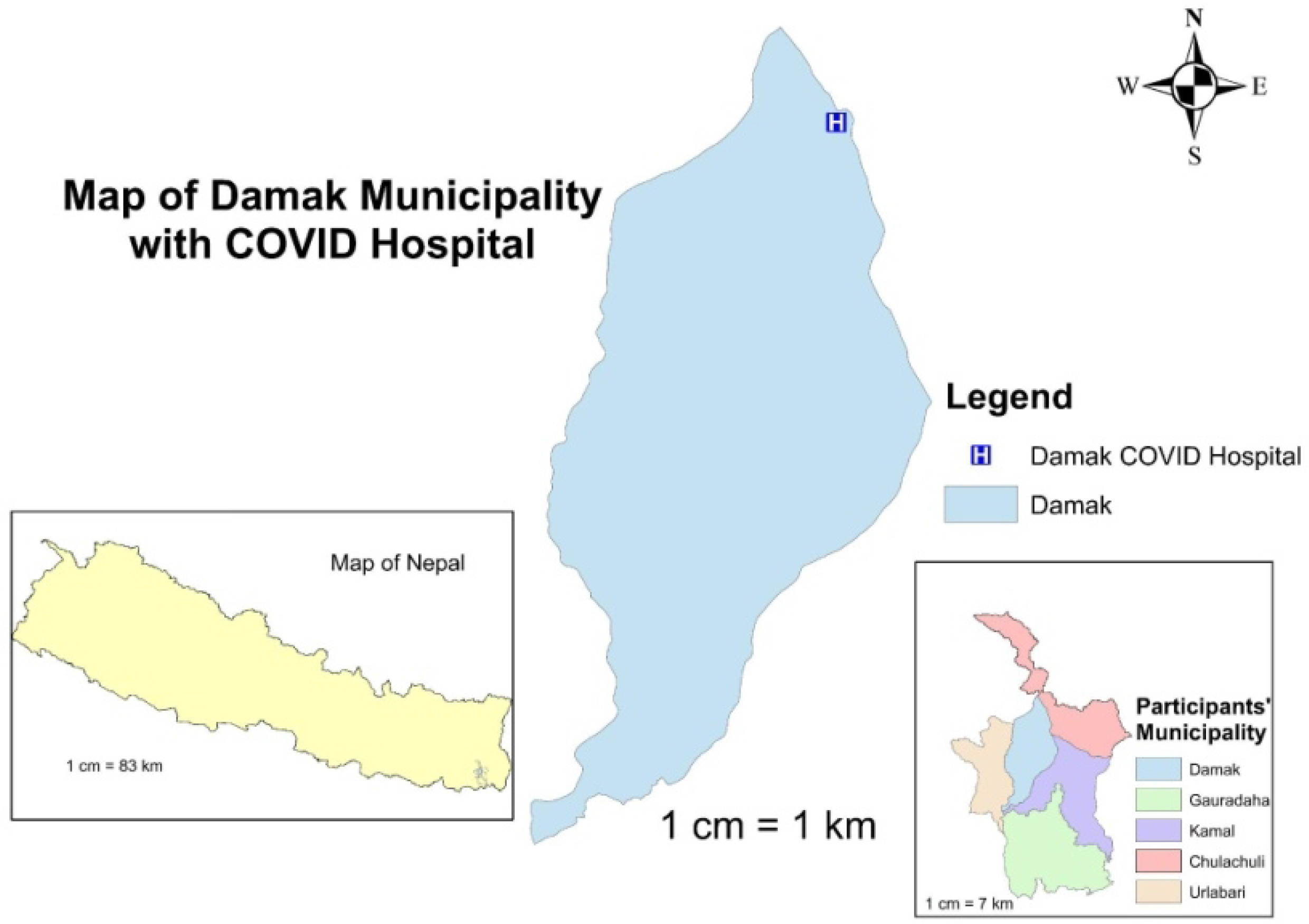
Map related to the study Area.

